# Voice of the patient: Emergence of new motor and non-motor symptoms in early Parkinson’s Disease?

**DOI:** 10.1101/2021.06.21.21258883

**Authors:** Michelle H.S. Tosin, Tanya Simuni, Glenn T. Stebbins, Jesse M. Cedarbaum

## Abstract

**Objective:** To explore the utility of using patient reported emergence of new symptoms (ES) as an outcome measure during the early phase of the disease.

**Methods:** We analyzed data from MDS-UPDRS Part IB and Part II from the Safety, Tolerability, and Efficacy Assessment of Isradipine for PD (STEADY-PD3) study, with at least one annual follow-up over two years. We divided the sample into categories of follow-up visit (between 0 and 12-months, and 13 and 24-months) and the number of ES for each part of the scale between participants who started symptomatic treatment and those who did not (STx-yes/no). We assessed ES differences between participants STx in each follow-up visit using Mann-Whitney U test, and the Kaplan-Meier analyses.

**Results:** Of 331 participants observed for months 0 to 12, 288 (87%) developed ES, and 182 (55%) started STx. For Part IB, the median number of ES did not significantly differ between the STx groups (Z=-0.86, p = 0.39), while for Part 2, the number of ES was significantly higher for the STx-yes group (Z=-2.38, p=0.02). Of 148 participants who continued to be observed for months 13 to 24, 114 (77%) developed ES, and 62 (42%) started STx. For Part IB, the median number of ES did not significantly differ between the STx groups (Z=-0.33, p = 0.74), while for Part 2, the number of ES was significantly higher for the STx-yes group (Z=-2.25, p=0.02).

**Conclusions:** Assessing ES among patient-reported experiences of daily living may provide a useful marker for tracking PD progression.

## Introduction

The Braak hypothesis holds that as Parkinson’s disease (PD) progresses, different areas of the brain become progressively invaded by the neurodegenerative process which manifests in behavioral changes ^1^. Although the clinical impact of PD is physically and visually obvious to most patients in the early stages of the disease, we do not have sensitive tools to assess disease progression specifically in early PD ^2-4^. Currently, to assess progression we rely mainly on observations of symptoms and functionality, measured with clinician completed scales and patient self-report measures ^5,6^. The progression of functional impairment over the course of the disease, especially in its earliest stages, seems almost imperceptible as measured by the current scales. However, in daily practice, clinicians and researchers are commonly struck by patient statements such as: “Last time I saw you I could do “X”, but now I can’t (or I need help, or it takes me longer)”. Disease progression, as viewed through this patient-centric lens of ever-accumulating milestones of difficulty to the point of failure, is not a linear process, but a stepwise, saltatory decline, with emerging symptoms (ES) or impairments piling on the old, one after another ^7,8^.

Measuring the impact of therapies designed to slow disease progression is thus rendered extremely challenging, with attempts from clinical trials to assess the clinical meaningfulness and statistical significance of interventions that might reduce by 30-50% an average disease progression rate of 5% per year. In this sense, determining how the measurement ES can outline the course of the disease, especially in patients with PD at an early stage, will contribute to the development of new health technologies based on patient centered outcomes ^9,10^.

A similar initiative in patients with early Alzheimer’s disease tracked the appearance of new neuropsychiatric symptoms, suggesting clinical relevance when associated with increased morbidity ^11^. In our study, we aimed to explore the utility of assessing ES impacting the daily experiences of patients with early PD, as measured by the Movement Disorder Society-sponsored revision of the Unified Parkinson’s Disease Rating Scale (MDS-UPDRS) Part IB and II in relation to the initiation of antiparkinson therapy (STx), as a potentially novel patient relevant outcome measure during the early phase of the disease.

## Methods

We analyzed data from the Safety, Tolerability, and Efficacy Assessment of Isradipine for PD (STEADY-PD) study, a multicenter, randomized, parallel-group, double-blind, placebo-controlled trial (ClinicalTrials.gov: NCT02168842). The aims and methods of the STEADY-PD study have been published elsewhere ^12^, as well as results ^13^.

### Data and Sample

From the enrolled cohort of 336 participants of the STEADY-PD dataset, we limited the sample to PD participants with complete data from MDS-UPDRS Part IB (Non-motor Aspects of Experiences of Daily Living) and Part II (Motor Aspects of Experiences of Daily Living), and with at least one annual follow-up for two years, totaling 331 participants. Because the outcome of the STEADY-PD study showed no effect of the investigative agent, in our analysis we combined participants receiving both placebo and active treatment.

### Outcomes

Our primary outcome was ES for participants during the course of the study. To perform this analysis, we divided the sample into categories according to the period of the follow-up visit, initiation of antiparkinson treatment or not (STx-yes or STx-no) during the observation period, and the presence of ES. We used STx as a proxy of patient- and clinician-perceived disease progression. We analyzed these outcomes for each part of the scale.

We separated the follow-up visits in two distinct periods: the first observation period was when the participants were evaluated between baseline and 12 months, and the second observation period was when the participants were evaluated between 13 months and 24 months. To assign STx categories, we used the date when antiparkinson therapy was initiated (the visit day) and analyzed it according to the time interval between follow-up visits. For example, if the subject started antiparkinson therapy on day 105 of the study, he was allocated to the first observation period between baseline and 12 months to the “STx-yes” group for the entire interval. If a participant started antiparkinson therapy on day 400 of the study, he was allocated to the second treatment period between 13 and 24 months.

We defined ES as the occurrence of a new symptom between the beginning of each period and the follow-up visit. For example, participants who were scored as zero on any given item on the MDS-UPDRS at the baseline and had any score other than zero at 12 months, were classified as having an ES. Those who were scored zero at the baseline, zero at 13 months and any score different from zero at 24 months were classified as having ES in the period between 13 months and 24 months.

### Statistical Analyses

We used tables and histograms with distribution of frequencies, medians, and percentages to summarize the descriptive statistics. Comparison ES between participant starting antiparksonian therapy (STx-yes) and those not on antiparkinsonian therapy (STx-no) were conducted as binomial tests. Mann-Whitney U test was used to compare differences between the two groups of participants with or without ES, regardless of STx status. Survival distributions for ES vs non-ES groups were analyzed using Kaplan-Meier curves. Statistical significance was set at alpha < 0.05 and analyses were corrected for multiple comparisons, where appropriate, using a Bonferroni correction. Finally, we estimated required sample size to detect at least a 30% change in ES over a 12-month period. All statistical analyses were performed using SPSS® Statistics version 26 (IBM reference)

## Results

At baseline, the 331 STEADY-PD participants included in this study had a mean age of 62.4 years (± 9.0), with a preponderance of males (72%). The mean disease duration from diagnosis was 10 months (± 8.8), and the Hoehn and Yahr stage median score was 2 (ranging from 0 to 3). The mean total for the Motor Examination (Part III) of the MDS-UPDRS was 25.4 (SD 10.4). For the MDS-UPDRS Parts that were analyzed for this study (Parts IB and II) the means were 4.1 (SD 3.02) and 5.24 (SD 3.95).

Of 331 participants observed in the first treatment period, 288 (87%) developed ES and 182 (55.0%) were STx-yes (p=0.078). Of 149 participants in the second treatment period one patient had missing values. Of the 148 remaining participants in the second treatment period 114 (77%) developed ES, and 62 (42%) were STx-yes (p=0.058) (Table 1).

**Table 1.** Emergent symptoms in participants with or without antiparkinsonian therapy measured by MDS-UPDRS Parts IB and II according to the follow-up visit

| 0 to 12 Months (n of patients=331, 100%) |  |  |  |  |  |  |  |  |  |  |  |  |  |
| --- | --- | --- | --- | --- | --- | --- | --- | --- | --- | --- | --- | --- | --- |
| MDS-UPDRS | STx-Yes<br>(n of patients=182, 55.0%) |  |  |  | STx-No<br>(n of patients=149, 45.0%) |  |  |  | Total of<br>patients<br>with ES<br>(%) | Total of ES<br>(%) | Binomial<br>Test for<br>Patients<br>STx Yes vs.<br>No | Mann-Whitney Z-score<br>(Patients) | p-value<br>(Patients) |
|  | N of<br>Patients<br>(%) | N of ES | ES<br>Median<br>(Min-Max) | Average of<br>ES per<br>patient | N of<br>Patients<br>(%) | N of ES | ES<br>Median<br>(Min-Max) | Average of<br>ES per<br>patient |  |  |  |  |  |
| ES-Part IB | 108 (56.8) | 179 (56.3) | 1 (0-4) | 0.98 | 82 (43.2) | 139 (43.7) | 1 (0-6) | 0.93 | 190 (57.4) | 318 (100.0) | 0.069 | -0.86 | 0.389 |
| ES-Part II | 142 (56.8) | 386 (61.8) | 2 (0-7) | 2.12 | 108 (43.2) | 239 (38.2) | 2 (0-9) | 1.6 | 250 (75.5) | 625 (100.0) | 0.037 | -2.38 | 0.017 |
| ES-Parts IB and II | 162 (56.3) | 565 (59.9) | 3 (0-11) | 3.1 | 126 (43.8) | 378 (40.0) | 2 (0-12) | 2.54 | 288 (87.0) | 943 (100.0) | 0.039 | -2.19 | 0.028 |
| 13 to 24 Months (n of patients=148, 100%) |  |  |  |  |  |  |  |  |  |  |  |  |  |
| MDS-UPDRS | STx-Yes<br>(n=62, 41.9%) |  |  |  | STx-No<br>(n=86, 58.1%) |  |  |  | Total of<br>Patients<br>with ES | Total of ES<br>(%) | Binomial<br>Test | Mann-Whitney Z-score<br>(Patients) | p-value<br>(Patients) |
|  | N of<br>Patients<br>(%) | N of ES | ES<br>Median<br>(Min-Max) | Average of<br>ES per<br>patient | N of<br>Patients<br>(%) | N of ES | ES<br>Median<br>(Min-Max) | Average of<br>ES per<br>patient |  |  |  |  |  |
| ES-Part IB | 25 (40.3) | 37 (41.1) | 1 (0-5) | 0.60 | 37 (59.7) | 53 (58.9) | 1 (0-3) | 0.62 | 62 (41.9) | 90 (100.0) | 0.162 | -0.33 | 0.738 |
| ES-Part II | 46 (46.5) | 120 (53.3) | 2 (0-7) | 1.94 | 53 (53.5) | 105 (46.6) | 2 (0-6) | 1.22 | 99 (29.9) | 225 (100.0) | 0.547 | -2.25 | 0.024 |
| ES-Parts IB and II | 49 (43.0) | 157 (49.8) | 3 (0-12) | 2.53 | 65 (57.0) | 158 (50.2) | 2 (0-8) | 1.84 | 114 (77.0) | 315 (100.0) | 0.16 | -1.529 | 0.126 |

STx: Symptomatic treatment starting during the interval | ES: Emergent symptoms

Based on the period treatment subsamples, we assessed the number of ES reported per participant separately for MDS-UPDRS Parts IB, II and for Parts 1B and II combined. Of the seven symptoms assessed by Part IB of the MDS-UPDRS there was a median ES of 1 (range = 0 – 4) in the STx-yes subsample in first period with an average of ES per participant of 0.98. In the second period, the median remained at 1 for the STx-yes subsample, but the range of ES increased to 0 – 5 with an average of ES per participant of 0.60. There was no significant difference between the number of participants with ES on Part IB in the STx-yes and STx-no groups in both periods (p=0.069 on the first treatment period and p=0.162 on the second treatment period). Also, ES of the Part IB were not significantly different between the STx-yes and STx-no groups for either treatment period (Z=-0.86, p=0.39 on the first treatment period and Z=-0.33, p=0.74 on the second treatment period) (Table 1), (see Supplemental 1 A and D). Of the thirteen symptoms assessed by Part II of the MDS-UPDRS, we found a median ES of 2 (range 0 - 7) in the STx-yes subsample for both follow-up periods, with an average of ES per participant of 2.12 in the first treatment period and 1.94 in the second treatment period. There was a significant difference between the number of participants with ES in the STx-yes and STx-no groups in the first treatment period year (p=0.037) but not in the second treatment period (p=0.547). Most participants had 0 or 1 ES in both the STx-yes and STx-no groups. However, there was a significant difference in the prevalence of ES when the therapeutic groups were compared in the two periods (Z=-2.38, p=0.02 for the first treatment period and Z=-2.25, p=0.02 for the second treatment period) (Table 1), (see Supplemental 1 B and E).

When we considered all 20 items of Parts IB and II of the MDS-UPDRS combined, we found a median of 3 ES (range 0-11) in the STx-yes subsample in first period with an average of ES per participant of 3.1, and a median of 3 (range 0-12) in the second treatment period with an average of ES per participant of 2.53. There was significant difference between the number of ES in the STx-yes and STx-no groups only in the first treatment period (Z=-2.19, p=0.039). There was a significant preponderance of participants in the STx-no subsample with two ES in the first period (26.8%, Z=-1.53 p=0.028). In the second treatment period, participants with one ES prevailed and no significance was found between the therapeutic groups (Table 1), (see Supplemental 1 C and F).

Next, we analyzed the pattern of individual symptoms experienced by the groups of participants at baseline and in the two treatment periods (Figure 1), (see Supplemental 2). Part IB (Figure 1, A) and Part II (Figures 1, B and C) item-analyses demonstrated that participants in the STx-yes group had significantly more ES related to Freezing (p=0.014), Eating Tasks (p=0.019), Walking and Balance (p=0.045) and Doing Hobbies (p=0.028) compared to the STx-no group in the first treatment period. In the second treatment period participants in the STx-yes group had significantly more ES related to Speech (p=0.002), Hygiene (p=0.007), Saliva and Drooling (p=0.043) and Eating Tasks (p=0.045) compared to the STx-no group. It should be noted, however that since MDS-UPDRS was performed only at the beginning and end of each interval, we cannot relate occurrence of ES to need for medication in this analysis.

**Figure 1.**
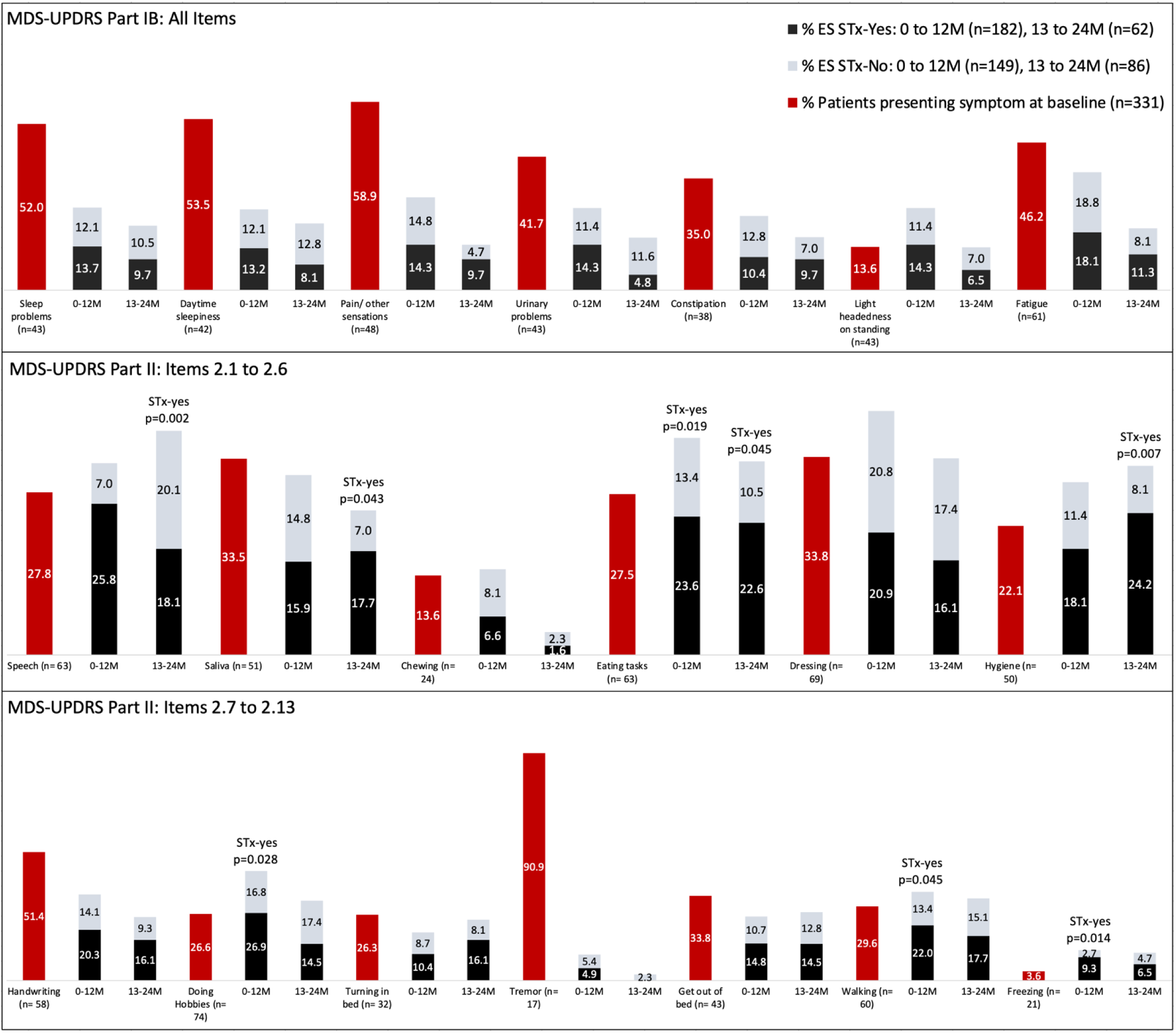
Proportion of participants endorsing individual MDS-UPDRS Part IB and II scale items at baseline and at follow-up study visits. Emergent symptoms (ES) reported at the follow-up timepoints are divided according to use of antiparkinson therapy (STx-yes and STx-no).

Given these results, we were interested to see how ES might perform as a clinical trial outcome measure. We estimate that a sample size of 98 would provide 0.80 power (1-β) to detect a 30% reduction in ES from baseline to 12-month follow-up, given an alpha of 0.05 and with equal assignment to treatment group and continuity correction when using Part IB alone. A sample size of 96 would be required for the same parameters when considering Part II alone. However, when both Parts IB and II are combined, a sample size of 82 would provide 0.80 power (1-β) to detect a 30% reduction in ES given the same parameters for alpha and subject assignment.

## Discussion

Kieburtz et al. recently suggested that tracking milestones of disease progression could provide a useful outcome measure for clinical trials of potential disease modifying therapies ^14^. However, milestones previously proposed, such as need for symptomatic medication, significant falls, or recognizable cognitive impairment, either represent changes in participant status relevant to more advanced disease or represent subjective and/or socially determined states. In early disease a milestone-based assessment of disease progression would of necessity need to be much more fine-grained.

In this exploratory analysis we asked whether, like the Braak progression of engagement of new brain areas concurrent with worsening of severity of pathology, the clinical progression of PD can be characterized by progressive appearance of ES, independent of the worsening severity of symptoms already present ^1,15^. Using data from Parts IB and 2 of the MDS-UPDRS in the STEADY-PD clinical trial ^12^, we found that the number of both motor and non-motor symptoms reported by participants increased over time in a clinical trial population, and that 87% of the study population reported at least one ES over the first 12 months of the study. Emergence of new motor symptoms was slightly more frequent than the emergence of non-motor symptoms, and the incidence, particular of motor ES was reduced in the group of participants who began STx during the first 12 months of the study. Thus, tracking self-reported ES may provide a novel means of assessing the progression of PD.

The results of our study demonstrated that Parts IB and II of the MDS-UPDRS, taken together as a single Patient-Reported Outcome (PRO) measure, can function as a record of milestone attainment in the form of appearance of new disease manifestations ^16^. Arguably, especially early in disease, appearance of a new symptom, as occurred in 87% of our participants within the first year of observation, could be interpreted to represent a significant milestone for most persons suffering from PD. The sensitivity of tracking ES as an outcome measure is reflected by the sample size estimates that less than 100 participants/arm would be required to observe a statistically significant effect in a 1-year clinical trial.

Our work does have limitations. First, our observation is based on a study in which the MDS-UPDRS was administered only at yearly intervals. At this point in time, data are not available in the public domain from other clinical studies that have administered the MDS-UPDRS more frequently than once every 6 or 12 months. Thus, we were unable, for example to assess the stability of ES once recorded. For ES to constitute a truly useful outcome measure, one would like to be able to verify stability of ES with observations at consecutive timepoints at least a month apart. Thus, it would be highly desirable to replicate our observations in a database with more-frequent MDS-UPDRS administration. Secondly, the clinical meaningfulness for participants of ES based on the MDS-UPDRS item inventory, while an attractive concept, has yet to be verified. Such verification could come either via the traditional scale validation and clinimetric methodology-use of Delphi panels, cognitive debriefing, and revalidation, or via correlation with patient self-reported experiences using approaches, such as the Patient Report of Problems (PROP) proposed by Vinikoor-Ilmer et al. based on data in the Fox Insight database^17^.

Finally, we found it interesting to note that the appearance of ES was slightly less frequent in study participants, all of whom were naïve to dopaminergic medications at enrollment, who began to receive STx during the study. Based on the available data it cannot be determined whether either a) STx delayed the onset of ES; b) Participants who started EX paradoxically had less ES during the time interval or c) STx masked the severity of ES that were present sufficiently to render them unremarkable using the MDS-UPDRS definitions. This topic could be a subject for further study. However, as has been reported, initiation of STx is the result of a complex medical and often social and economic calculus for individual participants, and factors like social circumstances, continuation of employment etc., may be more powerful determinants of STx initiation than emergence of any one or combination of symptoms ^18^.

## Conclusions

New symptoms continue to appear in most PD participants in the first 2 years of PD. Motor ES (Part II) were more frequent than non-motor ES (Part IB) among participants initiating antiparkinsonian treatment in both 0-12 and 13-24 months of the study. Assessing ES among patient-reported experiences of daily living may provide a useful marker for tracking PD progression. The concept of tracking ES as a clinical trial outcome measure is worthy of exploration in future studies and alternative datasets.

## Data Availability

Data are available on NINDS clinical trials repository
Archived Clinical Research Datasets | National Institute of Neurological Disorders and Stroke (nih.gov)
https://c-path.org/programs/cpp/
https://amp-pd.org/whole-genome-data

https://www.ninds.nih.gov/sites/default/files/sig_form_revised_508c.pdf

## Author Roles

1. Research project: A. Conception, B. Organization, C. Execution.
2. Statistical Analysis: A. Design, B. Execution, C. Review and Critique.
3. Manuscript Preparation: A. Writing of the first draft, B. Review and Critique.

M.H.S. Tosin: 1A, 1B, 1C, 2A, 2B, 2C, 3A.

T. Simuni: 3B.

G.T. Stebbins: 1A, 1B, 2A, 2C, 3B.

J.M. Cedarbaum: 1A, 1B, 2C, 3B.

## Disclosures

### 2. Financial Disclosures for the previous 12 months

**MHST** received grants and research from: Coordination for the Improvement of Higher Education Personnel (CAPES), International Parkinson and Movement Disorder Society. MHST Reports consulting with honoraria from Rush University Medical Center.

**TS** in the last 12 months has served as a consultant for Acadia, Caraway Therapeutics, Critical Path for Parkinson’s Consortium (CPP), Denali, General Electric (GE), Neuroderm, Sanofi, Sinopia, Sunovion, Roche, Takeda, MJFF and Voyager. Dr. Simuni served on the ad board for Acadia, Denali, General Electric (GE), Sunovion, Roche. Dr. Simuni has served as a member of the scientific advisory board of Caraway Therapeutics, Neuroderm and Sanofi. Dr. Simuni has received research funding from Biogen, Roche, Neuroderm, Sanofi, Sun Pharma, Amneal, Prevail, UCB, NINDS, MJFF, Parkinson’s Foundation

**GTS** reports consulting and advisory board membership with honoraria from: Acadia, Pharmaceuticals, Adamas Pharmaceuticals, Inc., Biogen, Inc., Ceregene, Inc., CHDI Management, Inc., Cleveland Clinic Foundation, Ingenix Pharmaceutical Services (i3 Research), MedGenesis Therapeutix, Inc., Neurocrine Biosciences, Inc., Pfizer, Inc., Tools-4-Patients, Ultragenyx, Inc., and the Sunshine Care Foundation. GTS received grants and research from: National Institutes of Health, Department of Defense, Michael J. Fox Foundation for Parkinson’s Research, Dystonia Coalition, CHDI, Cleveland Clinic Foundation, International Parkinson and Movement Disorder Society, and CBD Solutions. GTS reports honoraria from: International Parkinson and Movement Disorder Society, American Academy of Neurology, Michael J. Fox Foundation for Parkinson’s Research, Food and Drug Administration, National Institutes of Health, and the Alzheimer’s Association. GTS received salary from Rush University Medical Center.

**JMC** receives salary support from Coeruleus Clinical Sciences LLC and Yale Medical School. He has received honoraria from the Charcot Marie Tooth Research Foundation, the Michael J Fox Foundation, and the National Institutes of Health.

## Ethical Compliance Statement

We confirm that we have read the Journal’s position on issues involved in ethical publication and affirm that this work is consistent with those guidelines. This is a study of secondary data analysis and therefore did not need ethical approval.

