## Supplemental Data 1 for "Voice of the patient: Emergence of new motor and non-motor symptoms in early Parkinson’s Disease?"

Supplemental 1. Number of Emergent Symptoms in participants with or without antiparkinson therapy measured by MDS-UPDRS Parts IB and II according to the follow-up visit

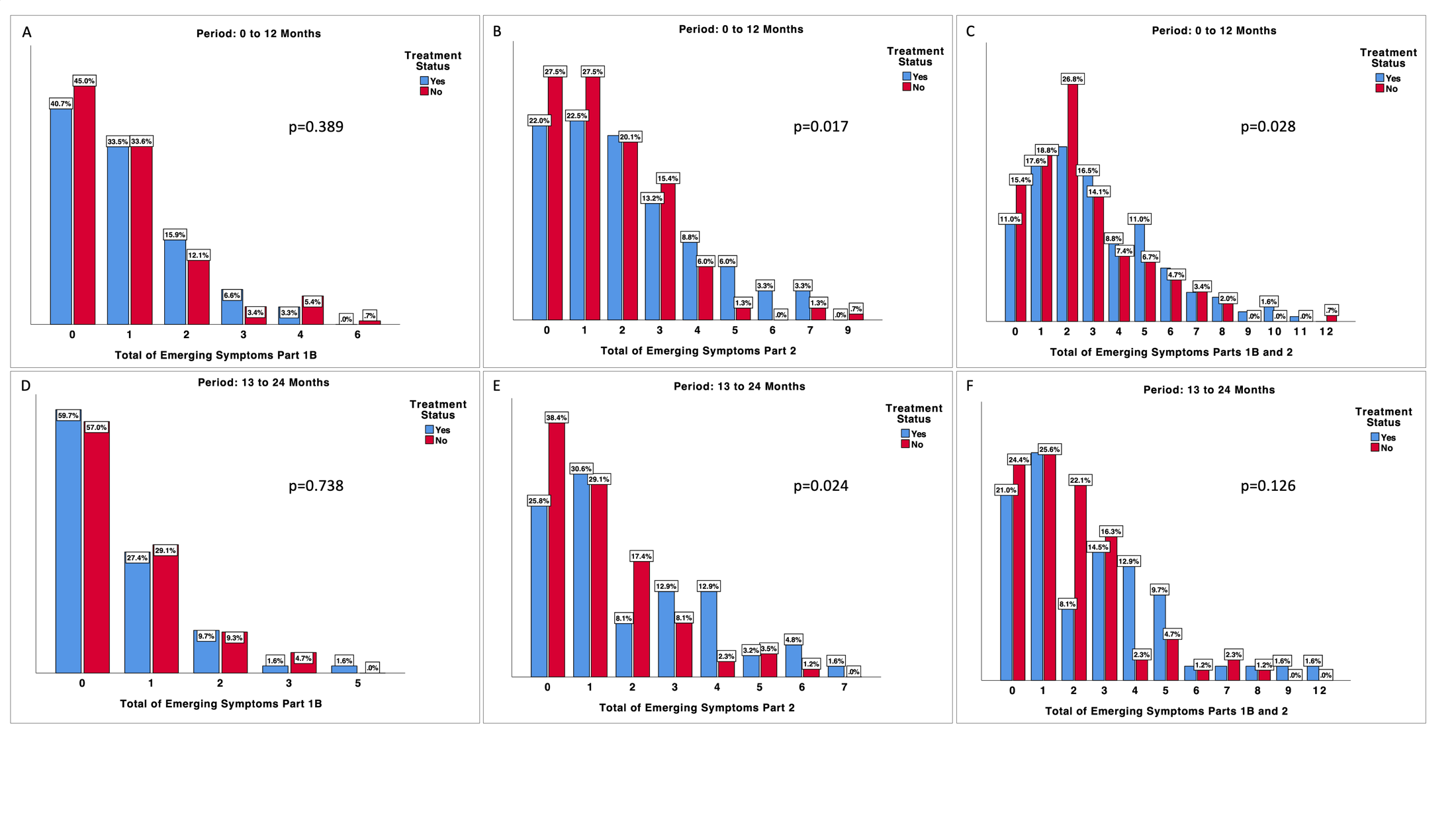
