## Supplemental Data 2 for "Voice of the patient: Emergence of new motor and non-motor symptoms in early Parkinson’s Disease?"

Supplemental 2. Distribution of Emergent Symptoms according to antiparkinsonian therapy and the follow-up visit

| Symptoms | Emergent Symptoms | 0 to 12 Months | | | | | | 13 to 24 Months | | | | | |
| --- | --- | --- | --- | --- | --- | --- | --- | --- | --- | --- | --- | --- | --- |
|  |  | Treatment Status | | | | Total (n=331) | | Treatment Status | | | | Total (n=148) | |
|  |  | Yes (n=182) | | No (n=149) | |  |  | Yes (n=62) | | No (n=86) | |  |  |
|  |  | N | % | N | % | N | % | N | % | N | % | N | % |
| Sleep problems | No | 157 | 54.5 | 131 | 45.5 | 288 | 87.0 | 56 | 42.1 | 77 | 57.9 | 133 | 89.9 |
|  | Yes | 25 | 58.1 | 18 | 41.9 | 43 | 13.0 | 6 | 40.0 | 9 | 60.0 | 15 | 10.1 |
| Daytime sleepiness | No | 158 | 54.7 | 131 | 45.3 | 289 | 87.3 | 57 | 43.2 | 75 | 56.8 | 132 | 89.2 |
|  | Yes | 24 | 57.1 | 18 | 42.9 | 42 | 12.7 | 5 | 31.3 | 11 | 68.8 | 16 | 10.8 |
| Pain and other sensations | No | 156 | 55.1 | 127 | 44.9 | 283 | 85.5 | 56 | 40.6 | 82 | 59.4 | 138 | 93.2 |
|  | Yes | 26 | 54.2 | 22 | 45.8 | 48 | 14.5 | 6 | 60.0 | 4 | 40.0 | 10 | 6.8 |
| Urinary problems | No | 156 | 54.2 | 132 | 45.8 | 288 | 87.0 | 59 | 43.7 | 76 | 56.3 | 135 | 91.2 |
|  | Yes | 26 | 60.5 | 17 | 39.5 | 43 | 13.0 | 3 | 23.1 | 10 | 76.9 | 13 | 8.8 |
| Constipation problems | No | 163 | 55.6 | 130 | 44.4 | 293 | 88.5 | 56 | 41.2 | 80 | 58.8 | 136 | 91.9 |
|  | Yes | 19 | 50.0 | 19 | 50.0 | 38 | 11.5 | 6 | 50.0 | 6 | 50.0 | 12 | 8.1 |
| Light headedness on standing | No | 156 | 54.2 | 132 | 45.8 | 288 | 87.0 | 58 | 42.0 | 80 | 58.0 | 138 | 93.2 |
|  | Yes | 26 | 60.5 | 17 | 39.5 | 43 | 13.0 | 4 | 40.0 | 6 | 60.0 | 10 | 6.8 |
| Fatigue | No | 149 | 55.2 | 121 | 44.8 | 270 | 81.6 | 55 | 41.0 | 79 | 59.0 | 134 | 90.5 |
|  | Yes | 33 | 54.1 | 28 | 45.9 | 61 | 18.4 | 7 | 50.0 | 7 | 50.0 | 14 | 9.5 |
| Speech | No | 149 | 55.6 | 119 | 44.4 | 268 | 81.0 | 46 | 36.5 | 80 | 63.5 | 126 | 85.1 |
|  | Yes | 33 | 52.4 | 30 | 47.6 | 63 | 19.0 | 16 | 72.7 | 6 | 27.3 | 22 | 14.9 |
| Saliva and drooling | No | 153 | 54.6 | 127 | 45.4 | 280 | 84.6 | 51 | 38.9 | 80 | 61.1 | 131 | 88.5 |
|  | Yes | 29 | 56.9 | 22 | 43.1 | 51 | 15.4 | 11 | 64.7 | 6 | 35.3 | 17 | 11.5 |
| Chewing and swallowing | No | 170 | 55.4 | 137 | 44.6 | 307 | 92.7 | 61 | 42.1 | 84 | 57.9 | 145 | 98.0 |
|  | Yes | 12 | 50.0 | 12 | 50.0 | 24 | 7.3 | 1 | 33.3 | 2 | 66.7 | 3 | 2.0 |
| Eating tasks | No | 139 | 51.9 | 129 | 48.1 | 268 | 81.0 | 48 | 38.4 | 77 | 61.6 | 125 | 84.5 |
|  | Yes | 43 | 68.3 | 20 | 31.7 | 63 | 19.0 | 14 | 60.9 | 9 | 39.1 | 23 | 15.5 |
| Dressing | No | 144 | 55.0 | 118 | 45.0 | 262 | 79.2 | 52 | 42.3 | 71 | 57.7 | 123 | 83.1 |
|  | Yes | 38 | 55.1 | 31 | 44.9 | 69 | 20.8 | 10 | 40.0 | 15 | 60.0 | 25 | 16.9 |
| Hygiene | No | 149 | 53.0 | 132 | 47.0 | 281 | 84.9 | 47 | 37.3 | 79 | 62.7 | 126 | 85.1 |
|  | Yes | 33 | 66.0 | 17 | 34.0 | 50 | 15.1 | 15 | 68.2 | 7 | 31.8 | 22 | 14.9 |
| Handwriting | No | 145 | 53.1 | 128 | 46.9 | 273 | 82.5 | 52 | 40.0 | 78 | 60.0 | 130 | 87.8 |
|  | Yes | 37 | 63.8 | 21 | 36.2 | 58 | 17.5 | 10 | 55.6 | 8 | 44.4 | 18 | 12.2 |
| Doing Hobbies and other activities | No | 133 | 51.8 | 124 | 48.2 | 257 | 77.6 | 53 | 42.7 | 71 | 57.3 | 124 | 83.8 |
|  | Yes | 49 | 66.2 | 25 | 33.8 | 74 | 22.4 | 9 | 37.5 | 15 | 62.5 | 24 | 16.2 |
| Turning in bed | No | 163 | 54.5 | 136 | 45.5 | 299 | 90.3 | 52 | 39.7 | 79 | 60.3 | 131 | 88.5 |
|  | Yes | 19 | 59.4 | 13 | 40.6 | 32 | 9.7 | 10 | 58.8 | 7 | 41.2 | 17 | 11.5 |
| Tremor | No | 173 | 55.1 | 141 | 44.9 | 314 | 94.9 | 62 | 42.5 | 84 | 57.5 | 146 | 98.6 |
|  | Yes | 9 | 52.9 | 8 | 47.1 | 17 | 5.1 | 0 | 0.0 | 2 | 100.0 | 2 | 1.4 |
| Getting out of the bed | No | 155 | 53.8 | 133 | 46.2 | 288 | 87.0 | 53 | 41.4 | 75 | 58.6 | 128 | 86.5 |
|  | Yes | 27 | 62.8 | 16 | 37.2 | 43 | 13.0 | 9 | 45.0 | 11 | 55.0 | 20 | 13.5 |
| Walking and balance | No | 142 | 52.4 | 129 | 47.6 | 271 | 81.9 | 51 | 41.1 | 73 | 58.9 | 124 | 83.8 |
|  | Yes | 40 | 66.7 | 20 | 33.3 | 60 | 18.1 | 11 | 45.8 | 13 | 54.2 | 24 | 16.2 |
| Freezing | No | 165 | 53.2 | 145 | 46.8 | 310 | 93.7 | 58 | 41.4 | 82 | 58.6 | 140 | 94.6 |
|  | Yes | 17 | 81.0 | 4 | 19.0 | 21 | 6.3 | 4 | 50.0 | 4 | 50.0 | 8 | 5.4 |
| Total | No | 3075 | 84.5 | 2602 | 87.3 | 5677 | 85.8 | 1083 | 40.9 | 1562 | 59.1 | 2645 | 89.4 |
|  | Yes | 565 | 15.5 | 378 | 12.7 | 943 | 14.2 | 157 | 49.8 | 158 | 50.2 | 315 | 10.6 |
